# Exposure Records Preserved by Two Users of Japan’s COVID-19 Contact-Confirming Application During the Seventh Wave: A Descriptive Study

**DOI:** 10.64898/2026.05.06.26352506

**Authors:** Shin-ichi Nakagawa, Seiji Kumagai, Akira Yamamoto

**Affiliations:** Research Institute of Info-Communication Medicine (RinCOM), Koganei, Tokyo, JPN; Faculty of Health Data Science, Juntendo University, Urayasu, Chiba, JPN

**Keywords:** COVID-19, digital contact tracing, exposure notification, contact tracing app, mandatory case reporting, Japan

## Abstract

**Introduction:** Japan’s COVID-19 Contact-Confirming Application (COCOA) stopped functioning in November 2022, after which no new records were generated. We describe the exposure records preserved by two users during the seventh wave of COVID-19 in Japan and the changes around the revision of September 26, 2022, when nationwide mandatory reporting of individual cases was restricted to specified patient groups (hereafter, the revision of full mandatory case reporting).

**Methods:** We analyzed exposure windows (date, duration, and minimum attenuation) contained in files exported from the app. Observer 1 worked at a hospital in Tokyo and commuted almost exclusively by private car; Observer 2 lived in Kyoto Prefecture and spent most of the time at home. Seven-day rolling sums of exposure windows were compared descriptively with daily new case counts in each prefecture.

**Results:** During the common pre-revision period (April 3 to September 25, 2022; 176 days), Observer 1 recorded 358 exposure windows and Observer 2 recorded 261. The rolling sums rose in July and fell in September, as did new case counts (r^2^ = 0.806 and 0.396), but week-on-week changes corresponded weakly (r^2^ = 0.183 and 0.001). Of the windows in this period, 97.2% and 93.1% had a minimum attenuation of 65 dB or more, the bucket given the lowest weight in the notification decision. After the revision, Observer 1 recorded three windows in 27 days (45 in the preceding 27 days), and Observer 2 recorded none in 23 days (25 in the preceding 27 days). On both devices, the match count in the OS exposure-check log fell to almost zero during the staged rollout of the app update of April 2022, while the app’s own export continued to record windows, and counts for recent exposure dates increased in later exports.

**Conclusions:** The exposure-window records of two users rose and fell with the seventh wave, although the correspondence of week-to-week changes was limited. Recorded windows decreased after the revision, but the effects of the policy change, the supply of diagnosis keys, app usage, and operating-system updates cannot be separated. This descriptive report does not evaluate the preventive effectiveness of COCOA. It shows that indicator definitions, retrieval dates, and the history of software and policy changes are essential when records left on users’ devices are analyzed after the fact.

## 1. Introduction

Japan’s COVID-19 Contact-Confirming Application (COCOA) was released by the Ministry of Health, Labour and Welfare (MHLW) on June 19, 2020 [1]. Built on the Google-Apple Exposure Notification (GAEN) framework [2,3], it recorded proximity via Bluetooth and notified users when a person they had been near later registered as positive. Positive registration required a processing number, which was issued only after a case notification under the Infectious Diseases Control Law had been entered into HER-SYS, the national case-management system. By November 16, 2022, the app had been downloaded 41,287,054 times, and 3,694,068 positive registrations had been made [4].

The effectiveness of such apps has been studied through simulation. Omae et al. used a multi-agent simulation to show that the reduction in cases depended on the app usage rate, the rate at which contact conditions were met, the registration rate of positive users, and the self-isolation rate, and that these parameters could not be determined from the data then available [5]. The same question has been examined with a compartmental model [6]. For the NHS COVID-19 app in England and Wales, Wymant et al. estimated the epidemiological impact from operational data and reported that a change in detection settings in October 2020 increased the number of notifications per case approximately 2.9-fold [7]. Kendall et al. examined the first year of operation and documented further changes to the notification threshold [8]. What a user observes therefore depends not only on the epidemic but also on the configuration of the system.

The government’s summary report (published in February 2023 and supplemented in March 2023) estimated, from data collected by the final version of the app, that approximately 9.58 million notifications were generated between April and September 2022, and that 26.5% of users received a notification between April and November of that year. The report also presented a “positive-signal reception rate” that included signals below the notification threshold [4]. The quantity analyzed in this study is close to that measure.

On September 13, 2022, following the decision to revise full mandatory case reporting, the government announced that COCOA would be discontinued. The revision took effect nationwide on September 26, restricting the issuance of processing numbers to persons at high risk of severe disease [4,9]. The app stopped functioning in November 2022, and the discontinued version generated no new records. The exports used in this study were made before the app stopped; records that were not preserved at the time cannot be regenerated. Approximately 15% of respondents to the final user survey had used the export function on which this study relies [4].

Regarding the public health effect of such apps, one assessment concluded that they prevented many infections even under a decentralized design [17], whereas it was pointed out from the outset that a decentralized design limits the means by which health authorities can verify the accuracy of notifications after the fact [18]. In 2021, the World Health Organization (WHO) and the European Centre for Disease Prevention and Control (ECDC) published an indicator framework for evaluating effectiveness in terms of adoption and use, speed of notification, and barriers and facilitators [19].

In this study, we describe the exposure records preserved by two of the authors during the seventh wave and around the revision of full mandatory reporting. This study does not evaluate the epidemiological effectiveness of COCOA. Its aim is to present, on the basis of actual records, findings on four questions that arise when records left on users’ devices are analyzed after the fact: how to handle the meaning and version of an indicator; how to handle the retrieval date and retrospective updating of records; how to include the supply of input data, namely diagnosis keys, in an evaluation; and what a user-verifiable export requires.

An earlier version of this work, which also compared the sixth and seventh waves, was posted to the medRxiv preprint server in May 2026 [10]. That comparison is not repeated here because the comparability of the on-screen value between the two periods could not be assured, as explained in Sections 3.5 and 5.1. Preliminary observations were presented to the Subcommittee on Prevention and Containment of Large-Scale Infectious Diseases, Section II, Science Council of Japan, on October 23, 2022.

## 2. Methods

### 2.1 Overview of COCOA and terminology

Figure 1 shows the system overview of COCOA and where the records used in this study reside. A device running COCOA generates a random key each day (the daily key), broadcasts via Bluetooth an identifier that is derived from this key and changes at regular intervals, and receives the identifiers broadcast by nearby devices, storing them together with the attenuation at reception for a fixed period [2,3]. A user diagnosed as positive enters into the app a processing number, which is sent to the patient on the basis of the case notification filed by the physician; that user’s recent daily keys are then uploaded to a server as Diagnosis Keys and distributed to all devices. Each device matches the distributed Diagnosis Keys against the identifiers it holds, on the device itself. The result of matching is retained in two places and in different forms: the OS exposure-check log (keys checked and match count) and the app export (exposure windows, daily summaries, and decision settings). Of these, what the user sees on the app’s ordinary screens is a notification, issued when the decision threshold is exceeded; part of the OS log (keys checked and match count) can be viewed on the iOS settings screen, and the app export is obtained by the user’s own operation. This study preserves these two kinds of records from the devices of two observers and describes them alongside the daily new case counts of their prefectures (public data). Figures concerning the registrant side of the pathway (left part of Figure 1) are taken from MHLW publications.

**Figure 1:**
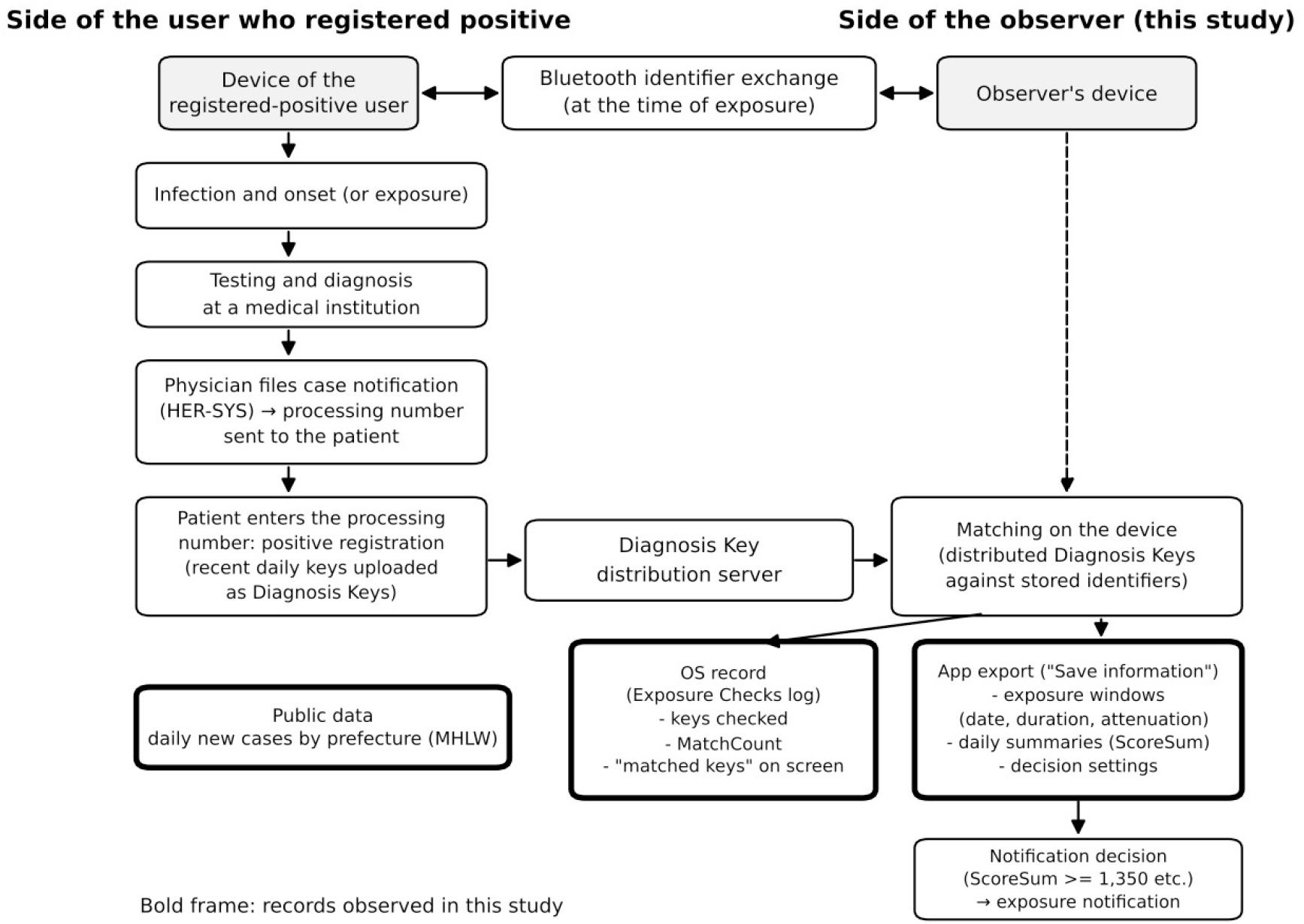
System overview of COCOA and the records used in this study. Bold frames indicate the records observed in this study.

From version 2.0.0, COCOA switched from version 1 to version 2 of the Exposure Notification application programming interface (API) [4,11]. Under version 1, the number of contacts meeting the conditions was counted; under version 2, reception time multiplied by a weight for each attenuation bucket is summed and compared with a threshold [11]. The design documentation describes the buckets as values that commonly occur in the following situations: 45 dB or less, very close proximity such as 0.5 m; 46-59 dB, 1.0 m between devices of the same type; 60-64 dB, 1.0 m between an iPhone and an Android device; and 65 dB or more, beyond 1.0 m for any combination of devices [11]. The terms used in this study are defined in Table 1.

**Table 1:**
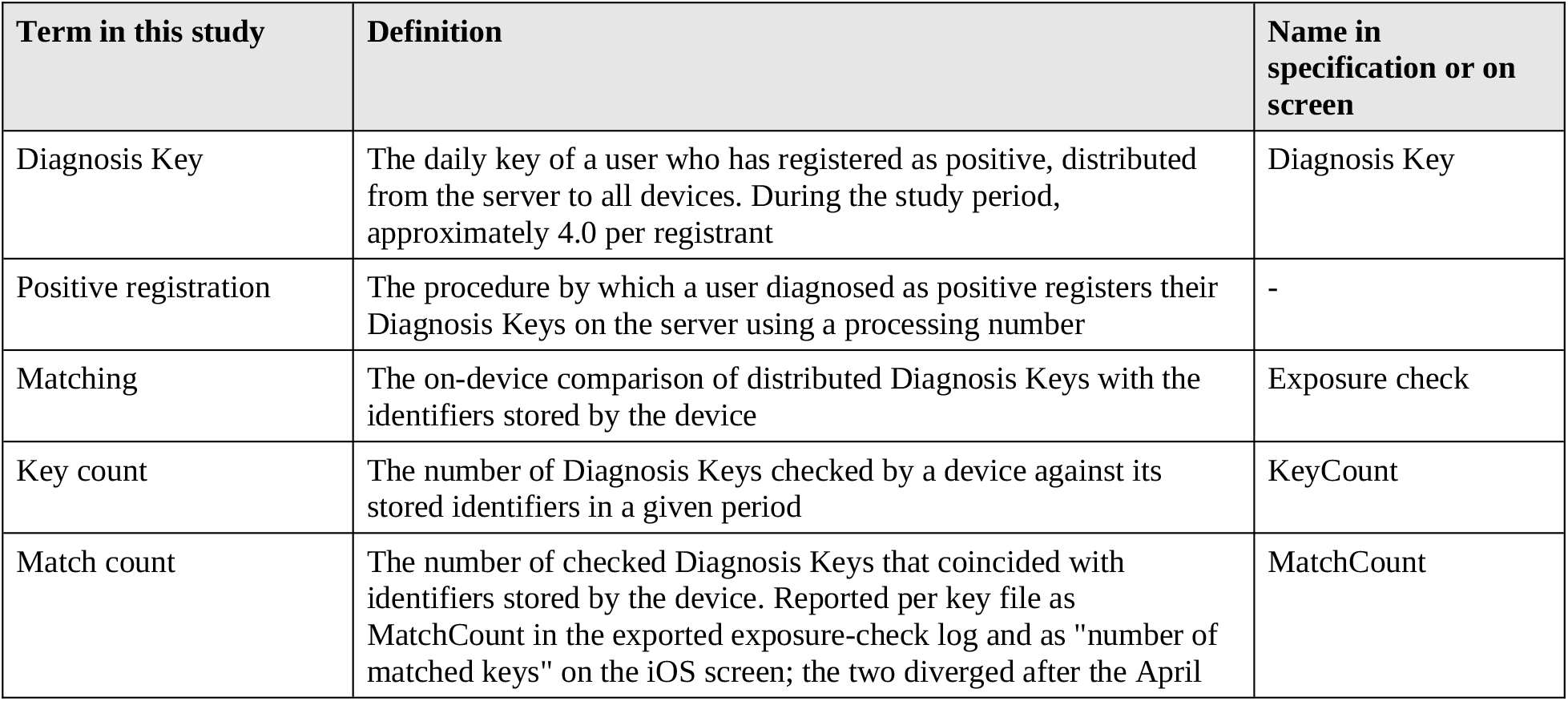
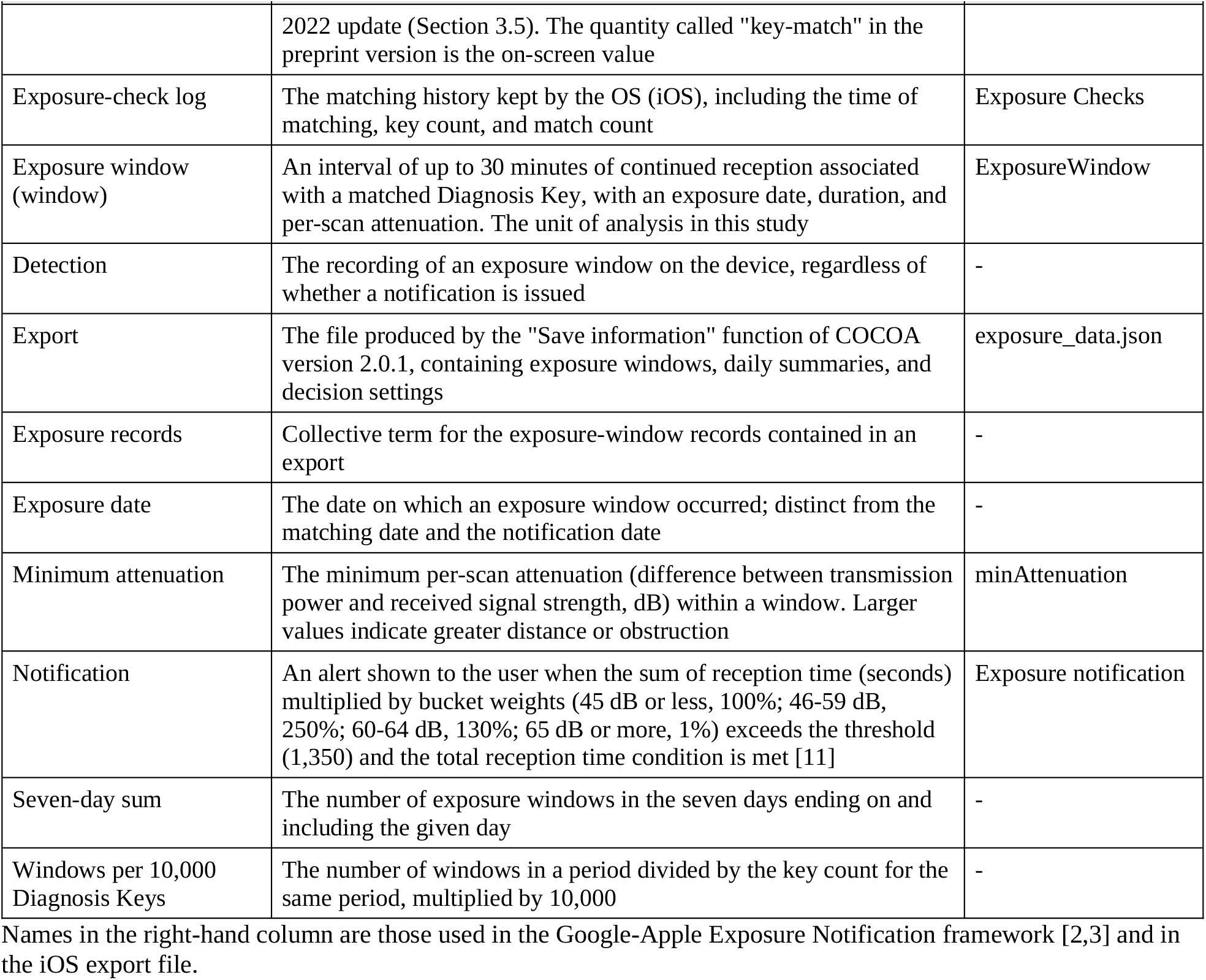
Terms used in this study.

| Term in this study | Definition | Name in specification or on screen |
| --- | --- | --- |
| Diagnosis Key | The daily key of a user who has registered as positive, distributed from the server to all devices. During the study period, approximately 4.0 per registrant | Diagnosis Key |
| Positive registration | The procedure by which a user diagnosed as positive registers their Diagnosis Keys on the server using a processing number | - |
| Matching | The on-device comparison of distributed Diagnosis Keys with the identifiers stored by the device | Exposure check |
| Key count | The number of Diagnosis Keys checked by a device against its stored identifiers in a given period | KeyCount |
| Match count | The number of checked Diagnosis Keys that coincided with identifiers stored by the device. Reported per key file as MatchCount in the exported exposure-check log and as "number of matched keys" on the iOS screen; the two diverged after the April | MatchCount |
|  | 2022 update (Section 3.5). The quantity called "key-match" in the preprint version is the on-screen value |  |
| Exposure-check log | The matching history kept by the OS (iOS), including the time of matching, key count, and match count | Exposure Checks |
| Exposure window (window) | An interval of up to 30 minutes of continued reception associated with a matched Diagnosis Key, with an exposure date, duration, and per-scan attenuation. The unit of analysis in this study | ExposureWindow |
| Detection | The recording of an exposure window on the device, regardless of whether a notification is issued | - |
| Export | The file produced by the "Save information" function of COCOA version 2.0.1, containing exposure windows, daily summaries, and decision settings | exposure_data.json |
| Exposure records | Collective term for the exposure-window records contained in an export | - |
| Exposure date | The date on which an exposure window occurred; distinct from the matching date and the notification date | - |
| Minimum attenuation | The minimum per-scan attenuation (difference between transmission power and received signal strength, dB) within a window. Larger values indicate greater distance or obstruction | minAttenuation |
| Notification | An alert shown to the user when the sum of reception time (seconds) multiplied by bucket weights (45 dB or less, 100%; 46-59 dB, 250%; 60-64 dB, 130%; 65 dB or more, 1%) exceeds the threshold (1,350) and the total reception time condition is met [11] | Exposure notification |
| Seven-day sum | The number of exposure windows in the seven days ending on and including the given day | - |
| Windows per 10,000 Diagnosis Keys | The number of windows in a period divided by the key count for the same period, multiplied by 10,000 | - |

Names in the right-hand column are those used in the Google-Apple Exposure Notification framework [2,3] and in the iOS export file.

### 2.2 Observers

Observer 1 (SN) is a physician. Throughout the study period, he left his home in Koganei, Tokyo, at 7:30 am by private car and drove approximately 25 km (about 60 minutes) to Akebono Hospital in Machida, Tokyo. There, he conducted appointment-based internal medicine outpatient clinics on Monday mornings and Thursday afternoons (20-26 patients per session) and worked at the health-screening center in the same building on other weekdays. On Saturday mornings, he worked at the health-screening center of Sayama General Clinic in Sayama, Saitama. On Sundays, he had breakfast at about 9:00 am at a family restaurant near his home. Apart from train trips to central Tokyo, he traveled exclusively by private car. His device (iPhone 12 mini; Apple Inc., Cupertino, CA, USA) was carried in his breast pocket, in a case whose material was not recorded, while commuting and at work; throughout the period he wore clothing with a breast pocket, including the white coat worn in the hospital. At home, the device was left on a desk to charge. He had been vaccinated against COVID-19 and did not contract the disease during the study period.

Observer 2 (SK) lives in Kyoto Prefecture. Throughout the study period, he kept outings to a minimum and spent most of the time at home. His device was an iPhone 12 Pro Max (Apple Inc., Cupertino, CA, USA); he carried it in a breast pocket when outside and left it on a desk when at home (self-reported). Whether a case was used was not recorded.

The daily routines of both observers remained broadly constant throughout the study period. The number of people around them, the movements of those people, and whether those people were running the app were outside the observers’ control.

### 2.3 Data and variable definitions

The unit of analysis is the exposure window (ExposureWindow) defined by the GAEN framework. In the public specification, an exposure window has a date, report type, infectiousness, calibration confidence, and a sequence of ScanInstances, each with a typical attenuation, a minimum attenuation, and the number of seconds since the previous scan [2]. One window represents up to 30 minutes of exposure, and longer exposures to the same Diagnosis Key are split into multiple windows [2]. In addition, each person who registered as positive uploaded multiple Diagnosis Keys, approximately 4.0 per registrant during the study period. A window therefore corresponds neither to one person nor to one encounter, and the counts reported below cannot be read as numbers of people.

Records were obtained from files produced by the “Save information” function of COCOA version 2.0.1. Each file contains the exposure windows held by the framework, daily summaries, and the app’s decision settings. The decision settings in the exports matched the attenuation buckets and weights described above, and treated the period from three days before to 10 days after symptom onset as infectious (the design document [11] gives two days before onset). Although the notification decision uses the mean attenuation of each scan [11], we classified windows by their minimum attenuation. In a window whose minimum attenuation is 65 dB or more, every scan falls in the bucket of 65 dB or more.

Observer 1 preserved several exports (with last exposure dates of August 5, August 16, August 21, and September 5, and one made in November before the app stopped functioning); the November export was used for the analysis. The recorded exposure dates range from March 29 to November 8, 2022 (109 exposure dates, 367 windows). The export date is not recorded in the file, but the OS version recorded in it (iOS 16.1.1) indicates that the export was made on or after November 10, the release date of that version. Observer 2 preserved several exports, the last on October 18, 2022 (exposure dates from April 3 to September 24; 74 exposure dates; 261 windows). Because exposure windows can be tabulated by exposure date, the notification date need not be used as a proxy for the exposure date. However, new case counts are generated through diagnosis and reporting and therefore have a different time base from exposure dates. Same-day comparisons in this study are descriptive and are not corrected for the lag between the two series.

The analysis periods and the rationale for treating days without records are shown in Table 2. For Observer 2’s device, the OS exposure-check log records a matching run on every day from April 3 to the final export date (October 18). This log shows that matching was performed; it does not guarantee that Bluetooth reception and the storage of records were normal throughout each day. Days without exposure windows in this period were treated as days on which matching was confirmed but no exposure window was recorded, and were counted as zero. The analysis period was extended to the export date, but, as described below, counts for roughly the 14 days before an export could increase later. To examine this, counts for the post-revision period excluding the final 14 days are also presented (Section 3.2, Table 4). For Observer 1, the OS exposure-check log survives for only 30 of the 208 days in the analysis period, so operation of the device cannot be confirmed from OS records. That he carried the device and kept the app running throughout the period is self-reported. Days without records were counted as zero, and the results depend on this treatment. The end of Observer 1’s analysis period (October 22) corresponds to the end of the Tokyo case series compiled at the time and was not planned in advance. He continued observing until the app was discontinued, and one window was recorded between October 23 and November 8.

**Table 2:** Summary of observers and records.

| Item | Observer 1 | Observer 2 |
| --- | --- | --- |
| Residence and activity | Tokyo (lives in Koganei; works at a hospital in Machida) | Kyoto Prefecture (spent most of the time at home) |
| Device | iPhone 12 mini | iPhone 12 Pro Max |
| OS (up to export) | iOS 15.6 to 16.1.1 | iOS 15.6.1 to 15.7 |
| App | COCOA 2.0.1 | COCOA 2.0.1 |
| Export date | On or after November 10, 2022 (not recorded) | October 18, 2022 (final) |
| Range of recorded exposure dates | March 29 to November 8, 2022 | April 3 to September 24, 2022 |
| Analysis period | March 29 to October 22 (208 days) | April 3 to October 18 (199 days) |
| Days with an exposure-check log entry | 30 | 199 |
| Treatment of days without records | Zero (operation self-reported) | Zero (matching confirmed on all days) |
| Days with exposure windows | 108 | 74 |
| Exposure windows | 366 | 261 |
| In common pre-revision period (April 3 to September 25) | 358 | 261 |
| Total duration | 813 min | 595 min |
| Minimum attenuation 45 dB or less | 0 (0.0%) | 0 (0.0%) |
| Minimum attenuation 46-59 dB | 8 (2.2%) | 5 (1.9%) |
| Minimum attenuation 60-64 dB | 2 (0.5%) | 13 (5.0%) |
| Minimum attenuation 65 dB or more | 356 (97.3%) | 243 (93.1%) |

Counts for Observer 1 refer to the analysis period (through October 22); one further window dated November 8 is not included. Days with an exposure-check log entry are days in the analysis period on which the OS exposure-check log records a matching run. Percentages are relative to each observer's total. OS: operating system.

Daily new case counts for Tokyo and Kyoto were obtained from the open data “Newly confirmed cases (daily)” published by the MHLW [20]. Key counts were obtained by day from the OS exposure-check log of Observer 2’s device; this log continues beyond the final app export and extends to November 16, 2022. Download counts, positive registrations, Diagnosis Key counts, and notification rates are taken from documents of the MHLW and the Digital Agency [4].

For Observer 1, the exported windows were also checked against the output of COCOAlog.jp, a public tool that analyzed the same export file and is mentioned in the government summary as having supplemented the app’s functions [4,12]. The output saved from the tool’s screen in mid-November 2022 matched the export file in the number of windows, total duration, and score for all 109 exposure dates. The output saved on August 17, 2022, showed 262 windows for March 29 to August 16 (75 exposure dates), fewer than the 270 windows for the same period in the November export. The eight additional windows were dated August 9 (one), 13 (three), 15 (one), and 16 (three), all within eight days before the August 17 output; counts for earlier exposure dates were identical in the two outputs. When the preserved exports were compared window by window, no window was lost in a later export, and windows added later to exposure dates already covered by an earlier export numbered 26 for Observer 1 and 5 for Observer 2, all dated within seven days before the last exposure date of the earlier export. For Observer 2, whose export dates are preserved in the file names, two, two, and one windows dated September 12 were added between the exports of September 14, 16, 17, and 19. For Observer 1, when the November export was compared with the last export before it (last exposure date September 5), no window had been added to any exposure date on or before September 5; the 26 windows mentioned above were added between successive exports, each within seven days of the preceding export’s last exposure date. The exports and the windows contained in each are listed in the Zenodo dataset (export_inventory.csv and exposure_windows_by_export.csv). Because the Diagnosis Keys of a registrant are distributed after the exposure date, counts for recent exposure dates can increase later. The site is no longer operational. This cross-check is reported to show that the values displayed by the tool at the time were a direct rendering of the export file; the analysis in this study does not depend on the tool.

A summary of the observers and records is given in Table 2.

### 2.4 Analysis

We computed backward seven-day sums of exposure windows. The period from April 3 to September 25, 2022 (176 days), in which the analysis periods of both observers overlap and which precedes the revision of full mandatory reporting, was defined as the common pre-revision period. Pearson correlations between each observer’s seven-day sums and the daily new case counts of the corresponding prefecture were computed for the 170 days from April 9 onward, when seven-day sums were available for both observers. Because seven-day sums overlap, these 170 points are not 170 independent observations. Values obtained with seven-day moving averages of new case counts are also reported. For Observer 1, correlations were also computed for the period through October 22.

For the comparison before and after the revision, the period from July 9, defined as the onset of the seventh wave, to September 25 (79 days) was compared with the period from September 26 onward. Because the earlier period contains the epidemic peak, a comparison between the 27 days immediately before the revision (August 30 to September 25) and the period after it is also presented. In addition, we calculated the number of new cases per window and, as a descriptive indicator of recorded windows relative to key count, the number of windows per 10,000 Diagnosis Keys. Key counts were deduplicated using the hash of each key file. Diagnosis Keys are tabulated by matching date and exposure windows by exposure date, and the two can differ by several days. Because all series are strongly autocorrelated, correlation coefficients are reported descriptively and are not used to infer effect sizes. To examine this point, we also computed correlations between week-on-week differences of non-overlapping weekly totals for the 25 full weeks from April 3 to September 24 (24 differences; the remaining day, September 25, was not used). The score used in the notification decision is recorded in the daily summaries (DaySummary) of the export as ScoreSum for each exposure date (reception time multiplied by the weight of each attenuation bucket and summed; Table 1), and this value was used here. Because a notification additionally requires a condition on total reception time in addition to ScoreSum reaching the threshold (1,350) [11], the notification decision cannot be reproduced from ScoreSum alone. The match counts that Observer 1 had written down from the iOS exposure-check screen (Section 3.5) were compared with the MatchCount in the exported log for the same dates through April 8 and, for July 9 to October 23, the last reading of each day was compared with the trailing 14-day sum of exposure windows in the November export ending on that day; days without a reading were omitted, and the comparison was repeated with the sum ending one day earlier or later and with the 7-day sum.

## 3. Results

### 3.1 Records of the two observers in the common period

During the common pre-revision period (April 3 to September 25, 2022; 176 days), Observer 1 recorded 358 exposure windows and Observer 2 recorded 261. The seven-day sums followed trends similar to the daily new case counts in each prefecture (Figures 2A, 2D). For the 170 points, the correlation was r = +0.898 (r^2^ = 0.806) for Observer 1 and Tokyo (Figure 2B) and r = +0.629 (r^2^ = 0.396) for Observer 2 and Kyoto (Figure 2E). With seven-day moving averages of new case counts, the corresponding values were r^2^ = 0.783 and 0.310. The correlation between the two observers’ seven-day sums was r = +0.720 (r^2^ = 0.519) (Figure 2C). In contrast, for week-on-week differences of weekly totals (24 differences), the correlation was r = +0.428 (r^2^ = 0.183) for Observer 1 and Tokyo and r = +0.035 (r^2^ = 0.001) for Observer 2 and Kyoto. Sharing a large epidemic peak is not the same as corresponding in week-to-week changes.

**Figure 2:**
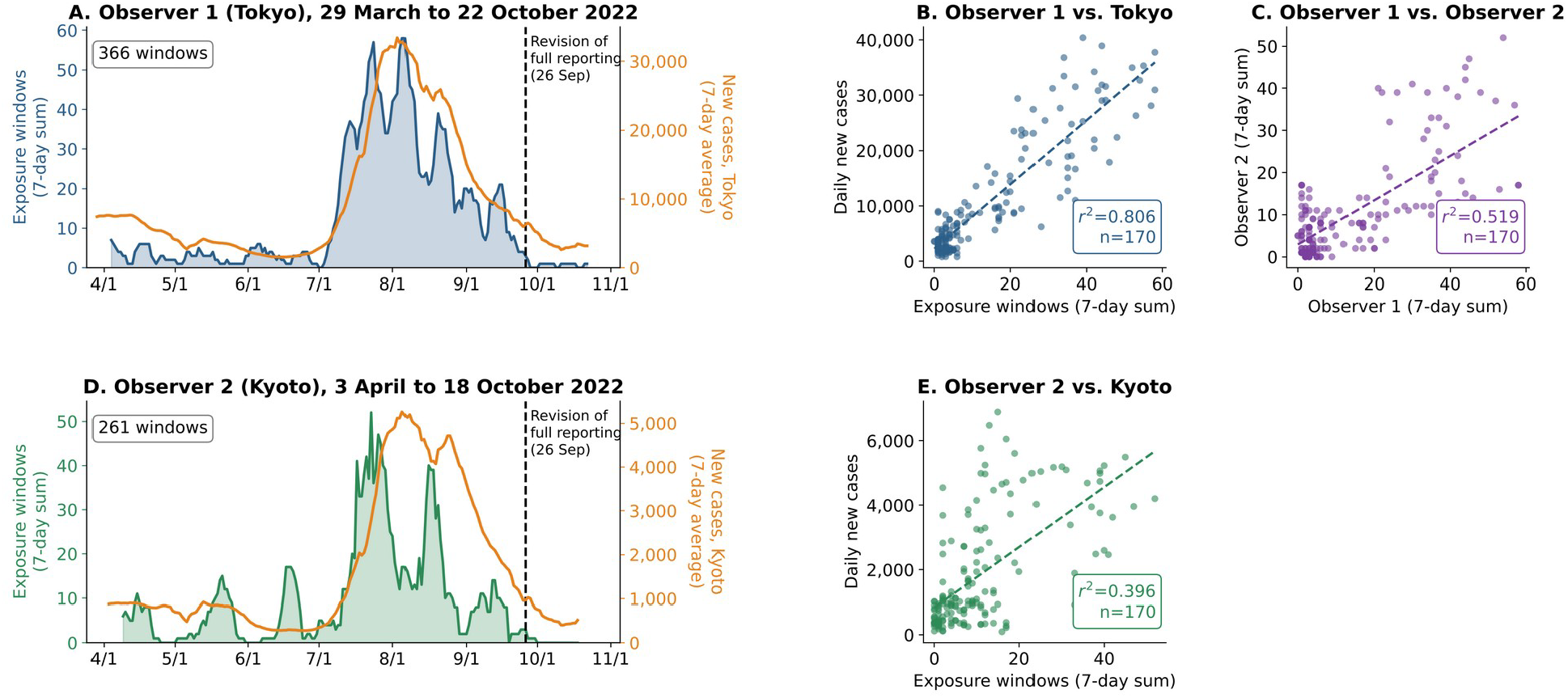
Exposure records of the two observers and new case counts in each prefecture (2022) (A) Observer 1 (Tokyo), March 29 to October 22. Blue, seven-day sum of exposure windows (left axis); orange, seven-day moving average of daily new cases in Tokyo (right axis). The dashed line marks September 26, when the revision of full mandatory reporting took effect nationwide. (B) Seven-day sums of Observer 1 versus daily new cases in Tokyo in the common pre-revision period (170 points; because seven-day sums overlap, these are not 170 independent observations). (C) Seven-day sums of the two observers against each other. (D) Observer 2 (Kyoto), April 3 to October 18. Green, seven-day sum of exposure windows; orange, seven-day moving average of daily new cases in Kyoto. (E) Seven-day sums of Observer 2 versus daily new cases in Kyoto. Dashed lines in B, C, and E are least-squares fits.

Monthly exposure windows per day and mean daily new cases in each prefecture are shown in Table 3. For both observers, exposure windows increased in July and decreased in September.

**Table 3:** Exposure windows per day, mean daily new cases, and windows per 10,000 Diagnosis Keys by month (common pre-revision period)

| Month | Observer 1 (Tokyo) |  |  | Observer 2 (Kyoto) |  |  |
| --- | --- | --- | --- | --- | --- | --- |
|  | Windows/day | New cases/day | Per 10,000 keys | Windows/day | New cases/day | Per 10,000 keys |
| April (from 3rd) | 0.43 | 6,038 | 0.156 | 0.57 | 823 | 0.208 |
| May | 0.39 | 3,252 | 0.097 | 0.71 | 720 | 0.178 |
| June | 0.37 | 1,967 | 0.162 | 0.70 | 312 | 0.310 |
| July | 4.77 | 18,314 | 0.427 | 3.52 | 2,459 | 0.315 |

|  | Observer 1 (Tokyo) |  |  | Observer 2 (Kyoto) |  |  |
| --- | --- | --- | --- | --- | --- | --- |
| Month | Windows/day | New cases/day | Per 10,000 keys | Windows/day | New cases/day | Per 10,000 keys |
| August | 4.42 | 24,488 | 0.251 | 2.23 | 4,472 | 0.127 |
| September (to 25th) | 1.52 | 8,700 | 0.259 | 0.96 | 1,839 | 0.164 |

Key counts were tabulated by day from the exposure-check log of Observer 2's device, summed over each month's comparison period, and used as a common value for both observers.

**Table 4:** Periods used in the comparison before and after the revision, with exposure windows, keys checked and new cases in each period.

| Period | Days | Observer 1 windows (per day) | Observer 2 windows (per day) | Keys checked (total) | Keys checked per day | Tokyo new cases per day | Kyoto new cases per day |
| --- | --- | --- | --- | --- | --- | --- | --- |
| July 9 to September 25 (before the revision) | 79 | 311 (3.94) | 196 (2.48) | 9,940,267 | 125,826 | 18,986 | 3,236 |
| August 30 to September 25 (27 days before) | 27 | 45 (1.67) | 25 (0.93) | 1,742,240 | 64,527 | 9,153 | 1,976 |
| September 26 to October 22 (after; Observer 1) | 27 | 3 (0.11) | — | 83,966 | 3,110 | 3,546 | 526 |
| September 26 to October 18 (after; Observer 2) | 23 | 2 (0.09) | 0 (0.00) | 76,003 | 3,304 | 3,564 | 536 |
| September 26 to October 4 (excluding the final 14 days) | 9 | 1 (0.11) | 0 (0.00) | 48,950 | 5,439 | 4,358 | 656 |

Key counts are from the OS exposure-check log of Observer 2's device (Section 2.3). The period through October 22 is the post-revision period of Observer 1; the period through October 18, and that period excluding the final 14 days, are those of Observer 2. One of Observer 1's three post-revision windows is dated October 19 or later. New cases are mean daily new cases.

For reference, the government survey reported that, among users of two years or longer, the number of notifications per person between April and September 2022 was 0.36 for those who did not commute, 0.44 for those who commuted by means other than public transport, and 1.31 for those who used trains or buses [4].

### 3.2 After the revision of full mandatory reporting

Observer 1 recorded 311 windows in the 79 days from July 9 to September 25 (3.94 per day) and three windows in the 27 days from September 26 to October 22 (0.11 per day) (Figure 2A). In the same comparison, mean daily new cases in Tokyo fell 5.4-fold, from 18,986 to 3,546, whereas exposure windows fell 35-fold. New cases per window rose from 4,823 to 31,915. Compared with the 27 days immediately before the revision (August 30 to September 25), windows fell 15-fold, from 45 (1.67 per day) to three, and mean daily new cases in Tokyo fell 2.6-fold, from 9,153 to 3,546. Observer 2 recorded 25 windows in the 27 days immediately before the revision and none in the 23 days from September 26 to the export date of October 18 (Figure 2D). In the same comparison, mean daily new cases in Kyoto fell 3.7-fold, from 1,976 to 536. Excluding the 14 days before the export, the count for September 26 to October 4 (nine days) was also zero. The end of Observer 1’s analysis period (October 22) is more than 14 days before his export (November 10 or later). The number of days, windows, keys checked and new cases in each period used in these comparisons are summarized in Table 4. Over the whole period from March 29 to October 22, the correlation between the seven-day sums of Observer 1 and daily new cases in Tokyo was r^2^ = 0.813 (202 points).

According to MHLW figures for the same period, positive registrations corresponded to 20.3%-23.8% of new cases in each week from July 1 to September 16, but to 1.4%-1.6% in each week from October 7 to 28. Diagnosis Keys per registrant remained approximately constant at about 4.0. Exposure windows are tabulated by exposure date, whereas the revision acted at the stage of positive registration. An exposure that occurred before the revision may remain unrecorded if the other person, after the revision, was no longer eligible to register and therefore could not upload Diagnosis Keys, so dividing the period at September 26 does not cleanly separate the periods before and after the revision.

### 3.3 Attenuation of recorded windows

The numbers of windows by minimum attenuation bucket are shown in Table 2 and Figure 3A. Windows with a minimum attenuation of 65 dB or more accounted for 356 of 366 (97.3%) for Observer 1 and 243 of 261 (93.1%) for Observer 2, and none had a minimum attenuation of 45 dB or less. Restricted to the common pre-revision period, the corresponding figures were 348 of 358 (97.2%) and 243 of 261 (93.1%). The weight of the bucket of 65 dB or more is 1% (Table 1), so reaching the notification threshold from this bucket alone would require approximately 37 hours of reception. Most recorded windows thus belonged to the attenuation bucket given a low weight in the notification decision. Because attenuation varies even at the same distance [11], however, the bucket does not represent actual distance or risk of infection. The ScoreSum recorded in the export for each exposure date reached the threshold (1,350) for Observer 1 only on August 4 (2,040; 13 windows, 124 minutes of reception); on all other days it was at most 240 (August 13), and for Observer 2 it was at most 360 (June 21). On Observer 1’s device, a screen saved on August 7 showed, for the contact of August 4, the app’s notice of “contact with a registered positive user” (119 minutes of contact in one day). Observer 1 thus received one notification during the study period, and its exposure date coincides with the only day on which the ScoreSum in the export reached the threshold. Observer 2 received no notification during the study period (self-reported). Restricting the seven-day sums to windows of 65 dB or more hardly changed the correlation with new case counts (170 points in the common pre-revision period; Observer 1, r^2^ = 0.806 for all windows and 0.801 for windows of 65 dB or more; Observer 2, 0.396 and 0.413, respectively).

**Figure 3:**
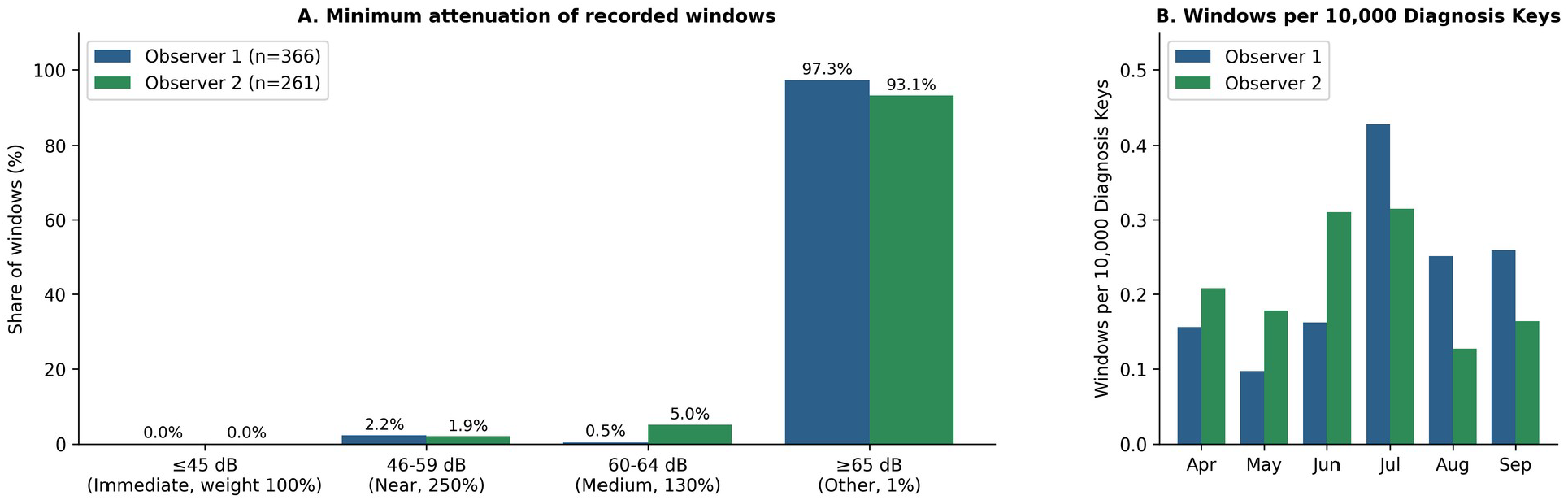
Attenuation of recorded windows and windows per 10,000 Diagnosis Keys. (A) Distribution of the minimum attenuation of recorded windows (whole analysis period). Buckets and weights follow the notification decision settings [11] (Table 1). (B) Windows per 10,000 Diagnosis Keys by month (common pre-revision period; Table 3).

Windows per 10,000 Diagnosis Keys by month (Table 3) are shown in Figure 3B.

### 3.4 Windows relative to key count

Diagnosis Keys are distributed nationwide, and in February and March 2022, when complete daily exposure-check logs were available for both devices, the key counts of the two devices were identical (1,949,549 keys each over the two months). Because the exposure-check log of Observer 1’s device is available only for some days from mid-April onward, key counts were derived by day from the log of Observer 2’s device and used as a common value for both observers. From April to September, windows per 10,000 Diagnosis Keys ranged from 0.097 to 0.427 for Observer 1 and from 0.127 to 0.315 for Observer 2 (Table 3, Figure 3B). Over the same period, mean monthly new cases varied approximately 13-fold in Tokyo and 14-fold in Kyoto. Across the revision, the key count fell from approximately 126,000 per day (July 9 to September 25) to approximately 3,100 per day (September 26 to October 22; approximately 65,000 per day in the 27 days immediately before the revision; Table 4), and windows per 10,000 Diagnosis Keys for Observer 1 were 0.313 and 0.357, respectively. With only three windows after the revision, however, this value does not show that detection performance was maintained. The indicator describes recorded windows relative to key count and is not adjusted for contact or infection risk.

### 3.5 Change in the quantity reported by the OS exposure-check log

Both observers also preserved the iOS exposure-check log, which records the key count and match count. For Observer 1, the match count was last non-zero on April 8, 2022; thereafter, no matches were reported among the 1.41 million Diagnosis Keys checked in the recorded period. For Observer 2, matches were reported intermittently until April 13, but from April 14 onward, among the 12.91 million Diagnosis Keys checked, the only match was one on May 21. Over the same period, the app’s own exports continued to record detections. The two devices were in different prefectures and were used differently. COCOA version 2.0.0, which supported version 2 of the Exposure Notification API, began to be distributed on April 7, 2022, in a staged rollout that expanded day by day [4]. The two dates, and the five-day interval between them, are consistent with the two devices having received this update on different days. The date on which each device received the update, however, cannot be confirmed from the records. The match count and the exposure windows therefore cannot be treated as a single series across this date, and we have not done so.

Separately, Observer 1 wrote down the “number of matched keys” shown on the iOS exposure-check screen, once a day from January 14 to April 20, 2022 (96 of 97 calendar days; no reading on March 23) and at each check from July 9 to October 23 (365 readings on 102 of 107 calendar days; no reading on July 16, 17, August 11, October 7, or 8). Through April 8, the total of the on-screen values (160) was almost identical to the total MatchCount in the exported log (161), and the weekly course of the two agreed. From April 9 to 12 the on-screen value was also zero, but from April 13 onward it remained non-zero while the exported MatchCount stayed at zero. From this date the on-screen value therefore no longer corresponded to the exported MatchCount. From July 9 to October 23, the on-screen value (last reading of each day, 102 days) moved with the 14-day trailing sum of exposure windows in the November export (r = 0.97; r = 0.83 against the 7-day sum), but it was not equal to that sum: it was lower on most days (median difference −6, interquartile range −15 to −1; equal on 8 of the 102 days and within ±2 on 32), the least-squares slope was 0.75 (intercept 1.9), and r was 0.98 with the sum ending one day earlier and 0.95 with the sum ending one day later. The on-screen value after the update was thus a quantity different from the exported MatchCount, and what it displayed cannot be identified from these records. The first version of this work [10] used these on-screen values.

## 4. Discussion

The exposure-window records preserved by the two users rose in July and fell in September during the seventh wave, in a manner similar to the new case counts of their prefectures. The correspondence of week-to-week changes, however, was weak, so the similarity reflects a shared large epidemic peak. In addition, exposure windows depend both on the supply of distributed Diagnosis Keys and on the contacts around the observer, and the former arises from the same process (diagnosis and registration) as the new case counts, so this shared process may contribute to the correlation between the seven-day sums and new cases. The number of windows per 10,000 Diagnosis Keys in Table 3 is a descriptive indicator normalized by the number of Diagnosis Keys checked. After the revision of full mandatory reporting, Observer 1 recorded three windows in 27 days and Observer 2 recorded none in 23 days, down from 45 and 25 windows, respectively, in the 27 days immediately before the revision. Over the same interval, new cases in Tokyo fell 2.6-fold and the key count fell approximately 21-fold, from about 65,000 to about 3,100 per day (Table 4). Positive registration in COCOA required a processing number linked to a case notification, and the revision restricted the issuance of these numbers. According to MHLW figures, positive registrations fell from more than 20% to less than 2% of new cases [4]. This sequence is consistent with the decrease in recorded windows, but the records in this study cannot separate the effects of the policy change, the decline of the epidemic, usage and registration behavior, and OS updates.

Most recorded windows belonged to the attenuation bucket given a low weight in the notification decision (65 dB or more), and the only notification during the study period was the one received by Observer 1 for the exposure of August 4. The government’s summary report likewise presented a positive-signal reception rate that included signals below the threshold [4]. Individual-level evaluations have been reported from Belgium [13], Switzerland [14], and Australia [15], each of which followed whether notifications reached contacts. Windows per 10,000 Diagnosis Keys ranged from 0.097 to 0.427 for Observer 1 and from 0.127 to 0.315 for Observer 2 between April and September, a narrower range than the approximately 13- to 14-fold variation in new cases over the same period. This is a descriptive indicator of recorded windows relative to key count, and the difference between the two observers may reflect individual behavior as well as differences in region and device; their contributions cannot be separated from these records.

The match count in the exported exposure-check log dropped to almost zero on the two devices five days apart in April 2022, during the staged rollout of COCOA version 2.0.0; thereafter, among 1.41 million and 12.91 million Diagnosis Keys checked, respectively, the only match was one for Observer 2. Over the same period, the app’s own exports continued to record detections, and on Observer 1’s device the match count shown on the screen was also non-zero (Section 3.5). Records of this kind can become discontinuous as a result of software updates.

### 4.1 Limitations

These records come from two individuals in two environments and cannot be generalized. Window counts cannot be converted into numbers of people, because registrants upload multiple Diagnosis Keys and exposures longer than 30 minutes are split into multiple windows. Because Observer 1 worked in Saitama Prefecture on Saturdays, one day per week is matched with case counts from a prefecture other than the one in which he spent that day. The OS of his device was iOS 15.6.1 on September 5, 2022, and iOS 16.1.1 at the time of the November export; with no records in between, the date of the update to iOS 16 cannot be determined. Because the revision (September 26) falls within this interval, a contribution of changed detection behavior to the post-revision decrease cannot be excluded. Windows per 10,000 Diagnosis Keys after the revision (0.357) were not lower than before it (0.313), but three windows are insufficient to judge whether detection behavior changed. Observer 2’s device ran iOS 15 throughout. No additional calibration or correction of the recorded values was applied by the authors. Weekly counts through September 25 decreased similarly for both observers, and no discontinuity specific to Observer 1 was seen in this period.

The post-revision period for Observer 2 is the 23 days up to the export date, and its end falls within the period whose counts could increase later. Exposure windows, new case counts, and key counts are based on exposure dates, dates of diagnosis and reporting, and matching dates, respectively, and the lags between them were not corrected. Because the revision acted at the stage of positive registration, its effect also extends to the end of the pre-revision period defined by exposure date. For Observer 1, counting days without records as zero relies on self-report. The match counts he wrote down from the screen are his own readings, and the date assigned to a reading may differ by about one day from the date of the check. The end of the analysis period and the start of the pre-revision comparison period (July 9) were not planned in advance. Because the exposure-check log of Observer 1’s device is available only for some days, key counts were taken from Observer 2’s device. Two other devices owned by Observer 1 recorded markedly fewer detections but were not analyzed, because their carrying conditions could not be reliably reconstructed. All series are autocorrelated, and the study period contains a single epidemic peak, so the correlation coefficients describe only the similarity of curve shapes. For week-on-week differences of weekly totals, the correspondence was weak for Observer 1 and absent for Observer 2. The post-revision period is short and contains only three windows, so its values lack precision. The sixth wave is outside the scope of this study: the app held exposure windows only from late March 2022, and the OS exposure-check log, which covers earlier periods, records a different quantity (the match count, not exposure windows; Section 3.5). For this reason, the comparison with the sixth wave made in the preprint version [10] is not repeated here.

## 5. Practical implications

The observations of this study and their implications for evaluating information systems are summarized in Table 5. The right-hand column contains proposals derived from the observations, not designs whose effectiveness was tested in this study.

**Table 5:** Observations in this study and implications for evaluating information systems.

| Observation or experience in this study | Implication for evaluating information systems |
| --- | --- |
| The match count in the exported OS log, the on-screen match count and the app exposure windows followed different courses across the update | Record the meaning and version of each log indicator, and do not join different indicators into one series |
| A later export contained more windows for past exposure dates | Manage retrieval dates, analysis cut-offs, and retrospective updates |
| The supply of Diagnosis Keys and the number of recorded windows decreased around the policy change | Include the supply process of input data, not only the users' logs, in the evaluation |
| Preserved exports allowed the operation of the system to be verified after the fact | Provide user-verifiable exports together with information on their provenance |

OS: operating system.

### 5.1 Points to note when handling records

Several issues arose in practice when handling the data.

First, both the OS exposure-check log and the app export should be preserved, with their roles kept distinct. In the first version of this work [10], the sixth and seventh waves were compared using the match counts that Observer 1 had written down from the OS screen. When the MatchCount in the exported exposure-check log and the on-screen values were later laid side by side for the same period, the two agreed through April 8 and diverged from April 13, and the on-screen values thereafter moved with, but were not equal to, the 14-day trailing sum of exposure windows in the export (Section 3.5). Because the comparability of the on-screen value between the sixth and seventh waves could not be assured, that comparison was withdrawn. In this study, on the other hand, the OS log was used to confirm operation (Section 2.3) and to derive key counts (Section 3.4), neither of which can be obtained from the export alone. Exports should be saved together with the settings they contain, and the OS log as the source of matching dates and key counts.

Second, the conditions under which a device is carried should be recorded. Two other devices owned by Observer 1 recorded markedly fewer detections, but their carrying conditions at the time could not be reliably reconstructed, and they could not be used in the analysis. In addition to the device model, OS version, and app version, the carrying position should be written down when recording begins.

Third, records indexed by exposure date remove the need to use the notification date as a proxy for the exposure date. The series with which they are compared (new case counts and key counts), however, each have their own time base, so it should be stated explicitly that the lag has not been corrected.

Fourth, the retrieval date of records must be managed. Counts for the same exposure dates differed between the output of August 17 and that of November; the eight windows added later were all dated within eight days before the August 17 output (August 9 to 16; Section 2.3). The retrieval date of each export should be recorded, and the analysis cut-off and the possibility that counts for recent days will increase should be stated. In this study, this was addressed by also presenting counts for the post-revision period excluding the final 14 days (Section 3.2).

For other users to carry out the same verification, the items to be preserved are: (a) the export file, including its settings, and its retrieval date; (b) the OS exposure-check log; (c) the device model, OS and app versions, and carrying conditions; and (d) the sources and access dates of the public series used for comparison. The data and the recalculation procedure used in this study are available on Zenodo (see Data availability).

### 5.2 Requirements for systems of this kind

Comparing these records with previous studies suggests several requirements for designing similar systems.

The first concerns the pathway for positive registration. Because registration in COCOA required a processing number linked to a case notification, positive registrations fell from more than 20% to less than 2% of new cases after the revision of full mandatory reporting [4]. Losses at the registration and notification stages have also been reported in other countries [13,14]. When the registration pathway depends on a notification system, the effect of changes to that system should be considered in advance.

The second concerns the continuity of measurement. In this study, the match count reported by the OS dropped to almost zero on both devices during the staged rollout of version 2.0.0 and diverged from the app’s own exports. In the United Kingdom, a change in detection settings was reported to increase notifications per case approximately 2.9-fold [7]. Because the quantities seen by users and evaluators depend on settings and versions, records should include the history of versions and settings, and it should be stated whether the same quantity is reported before and after each update.

The third concerns sub-threshold detections. In these records, most windows belonged to the bucket given a low weight in the notification decision; they were stored on the device but not shown to the user. In New South Wales, by contrast, 79 of 205 contacts detected by the app (39%) met the definition of a close contact [15]. Whether sub-threshold detections are retained, shown to users, or used for evaluation should be decided at the design stage.

The fourth concerns the preservation of records. Approximately 15% of respondents to the final survey had used the export function of COCOA [4], and records that users had not exported before the app was discontinued cannot be recovered. To allow verification after operations end, means for users to export and preserve records with their consent should be available while the system is in operation. That a decentralized design limits retrospective verification by the authorities was pointed out from the outset [18], and of the indicators called for in the WHO-ECDC framework [19], only some, such as the number of detections and its course over time, can be obtained from records on the user side.

All of these points concern the institutional pathways and settings on which a system depends. The Science Council of Japan has also recommended maintaining consistency with existing data when systems are modified and preserving the records of infectious disease countermeasures [16]. A simpler architecture that depends on fewer pathways and settings may be worth considering for systems of this kind.

The positive-registration pathway of COCOA depended on case notification. Registration pathways that do not depend on the notification system, such as authenticating test results by another route, remain to be examined. Proximity detection is meaningful for infections transmitted by droplets or through the air; which infections could be targeted, and whether such a system could help to suppress transmission, are also questions for future study.

## 6. Conclusions

The exposure-window records preserved by two COCOA users rose and fell in a manner similar to the seventh wave, although the correspondence of week-to-week changes was limited. Recorded windows decreased for both users after the revision of full mandatory reporting, but the effects of the policy change, the supply of Diagnosis Keys, app usage, and OS updates cannot be separated.

This study describes the records of two individuals; it neither evaluates the preventive effectiveness of COCOA nor shows how representative these records are of other users. We report them to document what a system that no longer exists recorded while it was operating, and to show that, when such records are analyzed after the fact, the definition of each indicator, the retrieval date, and the history of software and policy changes must be preserved together with the data.

If other users preserved exports from the same period, comparison with these records could show whether the phenomena described here were specific to these two environments. The authors are willing to take part in such comparisons.

## Data Availability

The exposure-window records, daily key-check counts, daily summary scores, the on-screen match counts written down by Observer 1, all preserved exports, derived daily series and a reproduction script are openly available on Zenodo (https://doi.org/10.5281/zenodo.22941040, version 3). Daily new case counts for Tokyo and Kyoto Prefecture were taken from the open data "Newly confirmed cases (daily)" published by the Ministry of Health, Labour and Welfare (https://covid19.mhlw.go.jp/public/opendata/newly_confirmed_cases_daily.csv). COCOA registration statistics are from the Digital Agency (https://www.digital.go.jp/policies/cocoa/).

https://doi.org/10.5281/zenodo.22941041

https://covid19.mhlw.go.jp/public/opendata/newly_confirmed_cases_daily.csv

## Additional information

### Author contributions

SN conceived the study, preserved and analyzed his own records, and drafted the manuscript. SK preserved his own records and contributed to data interpretation. AY contributed to study design, critical review of the analysis, and revision of the manuscript. All authors read and approved the final manuscript.

### Human subjects

The records analyzed are those of two of the authors’ own devices (SN, SK), both of whom consented to their use in research and publication. The records contain no information that could identify third parties. Ethics review: the study was not submitted to an ethics committee. Because it used only the authors’ own device records and publicly available aggregate case counts, and no third-party personal data were collected, the authors judged that review was not required.

### Data availability

Daily series derived from both observers’ exports, including an indicator of whether the OS exposure-check log contains an entry for each day, together with the match counts written down from the screen by Observer 1, the daily summary scores (ScoreSum) from the exports, the exposure windows in each preserved export, and the recalculation script (reproduce.py) that regenerates the principal figures in the manuscript, are provided as supplementary material (Zenodo, doi:10.5281/zenodo.22941040). New case counts for Tokyo and Kyoto are publicly available. The notification screen of Observer 1 saved on August 7, 2022, is available from the authors on request. The COCOAlog.jp outputs saved on August 17, 2022, and in mid-November 2022, and a copy of the pre-publication preview of the COCOA design document [11] saved on November 14, 2022 (https://gist.github.com/daisuke-nogami/808dfc8935cf4649e4272eddc13cdaf9), are available from the authors on request.

### Conflicts of interest

In compliance with the ICMJE uniform disclosure form, all authors declare the following. Payment/services info: All authors have declared that no financial support was received from any organization for the submitted work. Financial relationships: All authors have declared that they have no financial relationships at present or within the previous three years with any organizations that might have an interest in the submitted work. Other relationships: All authors have declared that there are no other relationships or activities that could appear to have influenced the submitted work.

## Acknowledgements

SN thanks his wife, Maki, whose support made it possible to keep the daily routine described here throughout the observation period. We thank Suminori Akiba, Midori Hirai, Hiroki Takakura, and Chihaya Koriyama of the Subcommittee on Prevention and Containment of Large-Scale Infectious Diseases, Section II, Science Council of Japan, for helpful comments, and Hiroaki Kitano (Okinawa Institute of Science and Technology) and Akimasa Hirata (Nagoya Institute of Technology) for their advice.

## Notes

### Competing Interest Statement

The authors have declared no competing interest.

### Author Declarations

The device records analyzed are the authors' own (S.N. and S.K.), who consented to their use in research and publication; no other individuals' data were collected, and the records contain no information that could identify third parties. No institutional ethics review was sought for the use of the authors' own records. All other data (prefectural case counts, COCOA statistics) are publicly available.

### Summary of Updates

Version 3 corrects one statement of fact in version 2 and extends the analysis of the preserved records. Version 2 stated that neither observer received a notification during the study period. A screen saved on 7 August 2022 shows that Observer 1 received one notification, for the exposure of 4 August 2022; this coincides with the only day on which the daily score recorded in the export reached the notification threshold. Sections 3.3 and 4 have been corrected accordingly. New case counts for both prefectures are now taken from the MHLW open data for the whole period, and the week-on-week comparison uses the 25 full weeks from 3 April to 24 September (24 differences); correlation coefficients changed in the third decimal place. The match counts that Observer 1 wrote down from the iOS screen, the daily summary scores from the exports, and a window-by-window comparison of all 14 preserved exports have been added (Sections 2.3, 2.4, 3.3 and 3.5). The account of the withdrawn sixth-wave comparison has been revised: the on-screen value and the exported match count diverged during the staged rollout of the April 2022 app update, and the comparability of the on-screen value between the two periods could not be assured. A system overview (Figure 1) and the carrying conditions of the devices have been added; the previous Figures 1 and 2 are now Figures 2 and 3. The companion dataset has been updated (Zenodo, version 3). Author information updated. This version has been submitted to a peer-reviewed journal.

